# ChatGPT in Healthcare: A Taxonomy and Systematic Review

**DOI:** 10.1101/2023.03.30.23287899

**Authors:** Jianning Li, Amin Dada, Jens Kleesiek, Jan Egger

## Abstract

The recent release of ChatGPT, a chat bot research project*/*product of natural language processing (NLP) by OpenAI, stirs up a sensation among both the general public and medical professionals, amassing a phenomenally large user base in a short time. This is a typical example of the ‘productization’ of cutting-edge technologies, which allows the general public without a technical background to gain firsthand experience in artificial intelligence (AI), similar to the AI hype created by AlphaGo (DeepMind Technologies, UK) and self-driving cars (Google, Tesla, etc.). However, it is crucial, especially for healthcare researchers, to remain prudent amidst the hype. This work provides a systematic review of existing publications on the use of ChatGPT in healthcare, elucidating the ‘status quo’ of ChatGPT in medical applications, for general readers, healthcare professionals as well as NLP scientists. The large biomedical literature database *PubMed* is used to retrieve published works on this topic using the keyword ‘ChatGPT’. An inclusion criterion and a taxonomy are further proposed to filter the search results and categorize the selected publications, respectively. It is found through the review that the current release of ChatGPT has achieved only moderate or ‘passing’ performance in a variety of tests, and is unreliable for actual clinical deployment, since it is not intended for clinical applications by design. We conclude that specialized NLP models trained on (bio)medical datasets still represent the right direction to pursue for critical clinical applications.

## 1 Introduction

In November 2022 a chat bot called ChatGPT was released. According to itself it is ‘a conversational AI language model developed by OpenAI. It uses deep learning techniques to generate human-like responses to natural language inputs. The model has been trained on a large dataset of text and has the ability to understand and generate text for a wide range of topics. ChatGPT can be used for various applications such as customer service, content creation, and language translation’. Since its release, ChatGPT has taken humans by storm and its user base is growing even faster than the current record holder TikTok, reaching 100 million users in just two months after its launch. ChatGPT is already used to generate textual context, presentations and even source code for all kinds of topics. But what does that mean specifically for the healthcare sector? What if the general public or medical professionals turn to ChatGPT for treatment decisions? To answer these questions, we will look at published works that already reported the usage of ChatGPT in the medical field. In doing so, we will explore and discuss ethical concerns when using ChatGPT, specifically within the healthcare sector (e.g., in clinical routines). We also identify specific action items that we believe have to be undertaken by creators and providers of chat bots to avoid catastrophic consequences that go far beyond letting a chat bot do someone’s homework. This review makes William B. Schwartz description from 1970 about conversational agents that will serve as consultants by enhancing the intellectual functions of physicians through interactions [94] as up-to-date as ever.

Even though the application of natural language processing (NLP) in healthcare is not new [34, 101, 111, 77], the recent release of ChatGPT, a direct product of NLP, still generated a hype in artificial intelligence (AI) and sparked a heated discussion about ChatGPT’s potential capability and pitfalls in healthcare, and attracted the attention of researchers from different medical specialities. The sensation could largely be attributed to ChatGPT’s barrier-free (browser-based) and user-friendly interface, allowing medical professionals and the general public without a technical background to easily communicate with the *Transformer* - and reinforcement learning-based language model. Currently, the interface is designed for question answering (QA), i.e., ChatGPT responds in texts to the questions*/*prompts from users. All established or potential applications of ChatGPT in different medical specialities and*/*or clinical scenarios hinge on the QA feature, distinguished only by how the prompts are formulated (Format-wise: open-ended, multiple choice, etc. Content-wise: radiology, parasitology, toxicology, diagnosis, medical education and consultation, etc.). Numerous publications featuring these applications have also been generated and indexed in *PubMed* since the release. This systematic review dives into these publications, aiming to elucidate the current state of employment, as well as the limitations and pitfalls of ChatGPT in healthcare, amidst the ChatGPT AI hype.

Based on the findings derived from existing publications on ChatGPT in healthcare, this systematic review addresses the following research questions:

- RQ1: What are the different medical applications where ChatGPT has already been tested?
- RQ2: What are the strengths, limitations and main concerns of ChatGPT for healthcare, especially with respect to the field they are applied to?
- RQ3: What are the key research gaps that are being investigated or should be investigated according to the existing works?
- RQ4: How can existing publications on ChatGPT in healthcare be categorized according to a taxonomy?

The rest of the manuscript is organized as follows: Section 2 briefly introduces NLP, transformers and large language Models (LLMs), on which ChatGPT is built. Section 3 introduces the inclusion criteria and taxonomy used in the systematic review, and discusses in detail the selected publications. Section 4 presents the answers to the above research questions (RQ1 - RQ4), and Section 5 summarizes and concludes the review.

## 2 Background

### 2.1 Natural Language Processing (NLP)

Natural Language Processing (NLP) [22] is an interdisciplinary research field that aims to develop algorithms for the computational understanding of written and spoken languages. Some of the most prominent applications include text classification, question answering, speech recognition, language translation, chat bots, and the generation or summarization of texts. Over the past decade, the progress of NLP has been accelerated by deep learning techniques, in conjunction with increasing hardware capabilities and the availability of massive text corpora. Given the fast growth of digital data and the growing need for automated language processing, NLP has become an indispensable technology in various industries, such as healthcare, finance, education, and marketing.

### 2.2 Transformer

In 2017, Vaswani et al. [109] introduced the Transformer model architecture, replacing previously widespread recurrent neural networks (RNN) [76], Long short-term memory networks (LSTM) [45] and Word2Vec [23]. Transformers are feedforward networks combined with specialized attention blocks that enable the model to attend to distinct segments of its input selectively. Attention blocks overcome two important limitations of RNNs. First, they enable Transformers to process input in parallel, whereas in RNNs each computation step depends on the previous one. Second, they allow Transformers to learn long-term dependencies. Since their introduction, Transformers consecutively achieved state-of-the-art results on various NLP benchmarks. Further developments include novel training tasks [24, 54, 114], adaptions of the network architecture [42, 64], and reduction of computational complexity [57, 64, 41]. However, the limited training data and the model complexities remained one of the primary factors of model performance. Transformers have also been used for tasks beyond NLP, such as image and video processing [95], and they are an active area of research in the deep learning community.

### 2.3 Large Language Models (LLMs)

Large language models (LLMs) [17] refer to massive Transformer models trained on extensive datasets. Substantial research has been conducted on scaling the size of Transformer models. The popular BERT model [26], which in 2019 achieved record-breaking performance on seven tasks in the Glue Benchmark [110], possesses 110 million parameters. On the other hand, GPT-3 [18] had already reached 175 billion parameters by 2021. At the same time, the size of the training datasets has continued to grow. BERT, for example, was trained on a dataset comprising of 3.3 billion words, while the recently published LLaMA [107] was trained on 1.4 trillion tokens. Despite the success of the LLMs, LLMs face several challenges, including the need for massive computational resources and the potential of adopting bias and misinformation from training data. Additionally, overconfidence when expressing wrong statements and a general lack of uncertainty remains to be a significant concern in NLP applications. As LLMs continue to improve and become more widespread, addressing these challenges and ensuring they are used ethically and responsibly is essential. ChatGPT is another representative LLM released by OpenAI and other tech giants have released their LLMs, such as the previously mentioned LLaMA from Meta, as a response. Figure 1 illustrates the evolution of LLMs.

**Figure 1:**
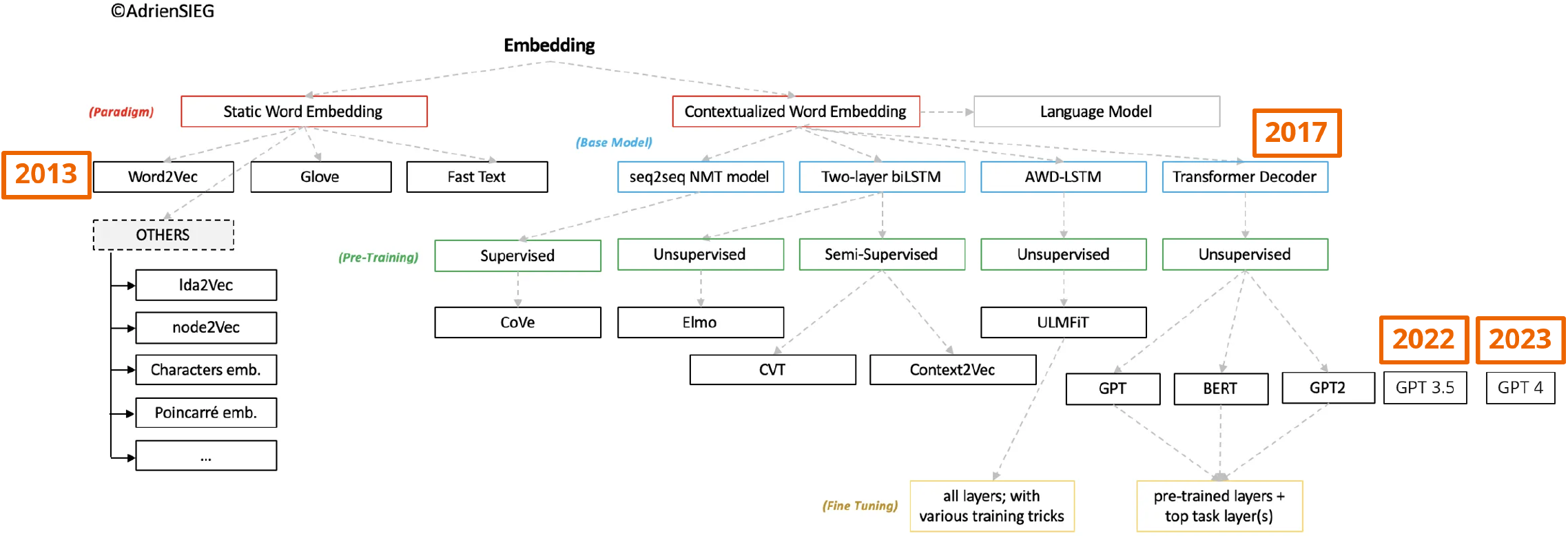
Evolution of large language models (LLMs) (adapted from [96]).

## 3 Methodology

The search strategy used in this systematic review is illustrated in Figure 2, following the PRISMA guidelines. We use *PubMed* as the only source to search candidate publications. Since the majority of the papers are very short (without abstracts), eligibility is determined at first screening based on the inclusion criteria below.

**Figure 2:**
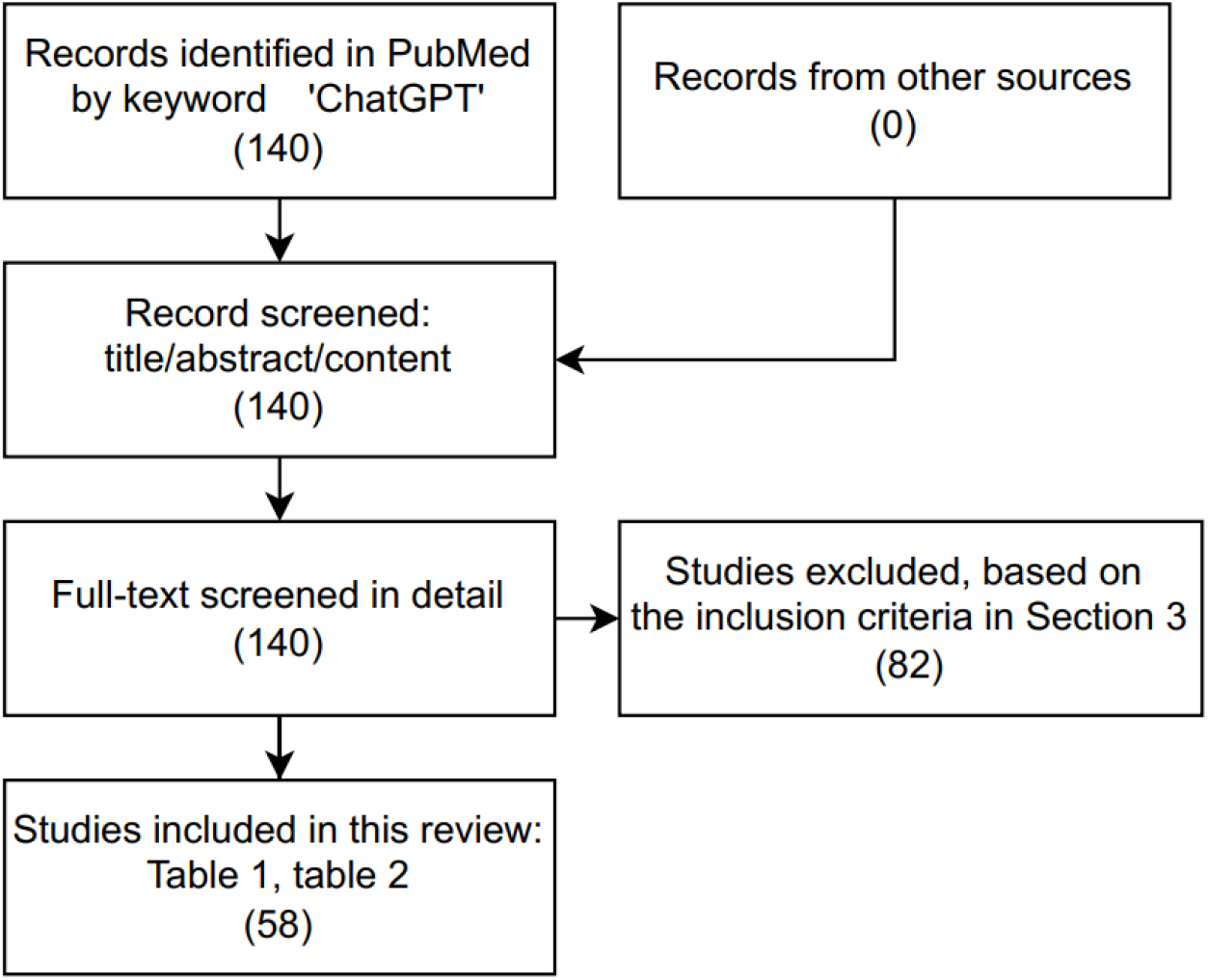
Search strategy used in this systematic review.

### 3.1 Inclusion Criteria

The review is expressly dedicated to the ChatGPT released in November 2022 by OpenAI, excluding its predecessors (*GPT-3*.*5, CPT-4*), other large language models (LLMs) such as *InstructGPT* and general NLP medical applications [69]. By March 20, 2023, a total of 140 publications are retrieved in *PubMed* (https://pubmed.ncbi.nlm.nih.gov/) using the keyword ChatGPT. Among them, article written in languages other than English (e.g., French [84]), without full text access (e.g., [62]), or whose main content has little to do with (or is not specific to) either ChatGPT (e.g., [46, 104, 33, 37]) or healthcare (e.g., [97, 103, 27, 6, 39, 13, 88, 21, 66, 115, 102, 43]) are excluded. Other representative exclusions include [44, 55], which deal with CPT-3, and [56, 30, 90, 2], where the authors claimed that ChatGPT assisted with the writing of the papers or case reports, but did not provide any discussion of the appropriateness of the generated texts and how the texts were incorporated into the main content. Generic comments that are not specific to healthcare, such as [105, 115, 16, 50], where the authors comment on the authorship of ChatGPT and using ChatGPT in scientific writing, are also excluded. Several repetitive articles were found from the *PubMed* search results. Table 1 and Table 2 show the full list of selected publications based on the inclusion (exclusion) criteria.

**Table 1:** Summary of *Level 1* and *Level 2* papers.

| Ref. | Scenario | Category | Main Content | Tag |
| --- | --- | --- | --- | --- |
| [93] | clinical workflow | editorial | discussion of the potential use, limitations and risks of ChatGPT in nursing practice | *Level 1* |
| [92] | medical research | perspective | comments about ChatGPT in scientific writing; Use ChatGPT to summarize and compare across papers | *Level 1* |
| [81] | medical research | editorial | generic comments on using ChatGPT in orthopaedic research | *Level 1* |
| [79] | medical research | letter to the editor | comments on using ChatGPT in scientific publications and generating research ideas | *Level 1* |
| [59] | miscellaneous | letter to the editor | comments on the potential use and pitfalls of ChatGPT in healthcare | *Level 1* |
| [106] | miscellaneous | editorial | discuss with ChatGPT about synthetic biology (e.g., applications, ethical regulations, history, research trends, etc.) | *Level 1* |
| [25] | medical research | editorial | comments on the pros and cons of using ChatGPT in medical research | *Level 1* |
| [65] | miscellaneous | original article | comments on the potential usage of ChatGPT in radiology (generate radiological reports, education, diagnostic decision-making, communicate with patients, compose radiological research article) | *Level 1* |
| [8] | medical education & research | letter to the editor | comments on the pros and cons of ChatGPT in medical education and research | *Level 1* |
| [35] | miscellaneous | primer | short comment on ChatGPT for Urologists | *Level 1* |
| [49] | consultation | correspondence | ChatGPT for antimicrobial consultation | *Level 1* |
| [48] | medical research | article (preprint) | comments on ChatGPT in peer-review | *Level 1* |
| [72] | miscellaneous | editorial | comments on ChatGPT in translational medicine | *Level 1* |
| [14] | consultation | letter to the editor | comments on the pros and cons of ChatGPT in public/community health (e.g., answer generic public health questions) | *Level 1* |
| [73] | miscellaneous | article | comments on the ethics of using ChatGPT in Health Professions Education | *Level 1* |
| [60] | medical research | letter to the editor | brief comments on using ChatGPT in medical writing | *Level 1* |
| [80] | medical education | editorial | comment on ChatGPT in nursing education | *Level 1* |
| [11] | miscellaneous | commentary | comment on ChatGPT in translational medicine | *Level 1* |
| [113] | miscellaneous | editorial | comment on ChatGPT in healthcare | *Level 1* |
| [58] | medical research | editorial | comment on ChatGPT in medical writing | *Level 1* |
| [7] | medical research | editorial | comment on using ChatGPT for scientific writing in sports & exercise medicine | *Level 1* |
| [12] | medical research | perspective | comment on medical writing | *Level 1* |
| [91] | miscellaneous | review | systematic review on ChatGPT in healthcare | *Level 1* |
| [5] | medical research | editorial | comment on the hallucination issue of ChatGPT in medical writing | *Level 1* |
| [71] | medical research | editorial | ChatGPT draft an article on vaccine effectiveness | *Level 2* |
| [108] | medical research | review | review on ChatGPT in medical research, including use examples | *Level 2* |
| [9] | medical research | original article | use ChatGPT to compile a review article on Digital Twin in healthcare | *Level 2* |
| [82] | clinical workflow | comment | use ChatGPT to generate a discharge summary for a patient who had hip replacement surgeries including follow-up care suggestions) | *Level 2* |
| [89] | clinical workflow | letter to the editor | ChatGPT gives diagnosis, prognosis and explanation for a clinical toxicology case of acute organophosphate poisoning | *Level 2* |
| [19] | medical research | editorial | ChatGPT answers questions about computational systems biology in stem cell research but its answers lack depth | *Level 2* |
| [40] | medical research | letter to the editor | use ChatGPT to search literature of a given topic, but majority of returned publications are fabricated | *Level 2* |
| [78] | medical (anatomy) education | letter to the editor | ChatGPT answers anatomy-related questions; Result shows ChatGPT is currently incapable of giving accurate anatomy information | *Level 2* |
| [1] | consultation | letter to the editor | ChatGPT answers questions on cardiopulmonary resuscitation | *Level 2* |
| [75] | miscellaneous | Discussions with Leaders (Invitation Only) | comment and use examples of ChatGPT in nuclear medicine | *Level 2* |
| [3] | medical education | editorial | ChatGPT answers multiple-choice questions on nuclear medicine; Results suggest ChatGPT does not possess the knowledge of a nuclear medicine physician | *Level 2* |
| [20] | medical research | brief report | comments on using ChatGPT in healthcare (e.g., compose medical notes) and medical research (e.g., generate abstracts, research topics) | *Level 2* |
| [47] | consultation | commentary | ChatGPT answers cancer-related questions information | *Level 2* |
| [15] | consultation | commentary | ChatGPT answersepilepsy-related questions | *Level 2* |
| [100] | consultation | article | comments on ChatGPT in diabetes self management and education (DSME) | *Level 2* |
| [31] | medical research | editorial | ChatGPT generates a curriculum about AI for medical students and a list of recommended readings) | *Level 2* |

**Table 2:** Summary of *Level 3* papers.

| Ref. | Scenario | Summary | Results/Conclusion | Version | Journal |
| --- | --- | --- | --- | --- | --- |
| [87] | clinical workflow | decide an imaging procedure or evaluate whether a procedure is proper for breast cancer/pain patients | specialized ChatGPT is needed | Jan. 9, 2023 | preprint |
| [86] | clinical workflow | ChatGPT supports clinical decision-making, by answering questions from Merck Sharpe & Dohme (MSD) clinical vignettes | ChatGPT achieves an overall accuracy of (71.7%) on 36 clinical vignettes covering the entire clinical workflow | Jan. 9, 2023 | preprint |
| [51] | medical education | compare ChatGPT with medical students in (an internal) parasitology exam (79 questions) | ChatGPT is not comparable to medical student (Acc. 89.6%) in parasitology questions | Dec. 15, 2022 | JEEHP |
| [36] | medical education | ChatGPT takes US Medical Licensing Examination (USMLE) | ChatGPT achieved passing score | Dec. 15, 2022 | JMIR |
| [4] | clinical workflow | ChatGPT writes patient letters (e.g., communicates diagnostic results, gives treatment advice) for 38 clinical scenarios | ChatGPT achieved high scores on both the factual correctness and humanness criterion | - | Lancet Digit Health |
| [99] | medical education | compare ChatGPT with medical students in parasitology exam (288 questions) from the Doctor of Veterinary Medicine (DVM) exam | ChatGPT and students achieve similar scores | - | Cell |
| [68] | clinical workflow | ChatGPT answers clinical decision support (CSD) alerts from Epic EHR | ChatGPT's answers are biased and redundant, their acceptability in CDS is low | - | preprint |
| [61] | medical education | ChatGPT takes USMLE (June 2022) | ChatGPT achieved passing score, and its explanations contain novel insights | - | PLOS Digital Health |
| [32] | medical education | ChatGPT takes life-support exams (AHA BLS / CLS Exams 2016) | ChatGPT did not reach passing score | Jan. 9 and 30, 2023 | Resuscitation |
| [53] | consultation | ChatGPT provides cancer-related information and feedback on cancer misconceptions | ChatGPT provides highly accurate cancer information | Dec. 15, 2022 | JNCI Cancer Spectrum |
| [67] | medical research | compared 50 ChatGPT-generated abstracts with real abstracts from scientific publications | Grammarly can detect ChatGPT-generated abstracts with high accuracy | - | AJOG |
| [83] | consultation | evaluate ChatGPT using 100 questions about retinal diseases | ChatGPT is highly accurate on general questions but less accurate for treatment options | - | Acta Ophthalmologica |
| [28] | consultation | compare ChatGPT with humans on 85 genetics/genomics questions | ChatGPT and humans perform similarly | - | preprint |
| [52] | consultation<br>medical education | ChatGPT answers 284 question from various medical specialties | ChatGPT achieved overall high accuracies | - | preprint |
| [112] | medical education | ChatGPT takes Chinese National Medical Licensing Examination | ChatGPT's performance on the exam is well below passing level | - | preprint |
| [63] | medical research | ChatGPT identifies research questions in gastroenterology (e.g., microbiome, endoscopy) | ChatGPT generates highly relevant but non-novel research questions | Dec. 15, 2022 | Scientific Reports |
| [38] | medical research | ChatGPT generates systematic review topics in plastic surgeries | ChatGPT performs moderately in generating novel systematic review ideas | - | Aesthetic Surgery Journal |
| [98] | consultation<br>medical education | evaluate ChatGPT using 100 OE questions about pathology | ChatGPT scored around 80% | Jan. 30, 2023 | Cureus |

### 3.2 Taxonomy

We propose a taxonomy, as shown in Figure 3, to categorize the selected publications included in the review. The taxonomy is based on applications, including ‘triage’, ‘translation’, ‘medical research’, ‘clinical workflow’, ‘medical education’, ‘consultation’, ‘multimodal’, each targeting one or multiple end-user groups, such as patients, healthcare professionals, researchers, medical students and teachers, etc. An application-based taxonomy allows more compact and inclusive grouping of papers, compared to categorizing papers by specific medical specialities. For example, scientific progress and findings generated through clinical practices are documented in the form of publications and*/*or reports, and literature reviews and novel ideas are usually required for medical researchers of all disciplines to publish their works. Thus, papers on ‘scientific writing’, ‘literature reviews’, ‘research ideas generation’, etc., can be grouped into the ‘medical research’ category. Similarly, the ‘consultation’ category comprises papers where ChatGPT is used in medical consulting settings for both corporations (e.g., insurance companies, medical consulting agencies, etc.) and individuals (e.g., patients) seeking medical information and advice. The ‘clinical workflow’ category includes ChatGPT’s applications in a variety of clinical scenarios, such as diagnostic decision-making, treatment and imaging procedure recommendation, and writing of discharge summary, patient letter and medical note. Furthermore, clinical departments, regardless of medical specialities, may benefit from a translation system for patients*/*visitors who are non-native language speakers (‘translation’). A triage system [10] guiding patients to the right departments would reduce the burden of clinical facilities and centers in general. Note that different categories are not necessarily completely independent, since all applications are reliant upon the QA-based interface of ChatGPT. By formulating the same questions differently according to different scenarios, ChatGPT’s role can change. For instance, reformulating multiple choice questions about a medical speciality in medical exams to open-ended questions, ChatGPT’s role changes from a medical student (‘medical education’) to a medical consultant (‘consultation’) or a clinician providing diagnosis or giving prescriptions (‘clinical workflow’). To avoid such ambiguity, categorization of a paper is solely based on the scenario explicitly reported in the paper. The connections between the applications and end-users in Figure 3 are also not unique. In this review, only the most obvious connections are established, such as ‘medical education’ - ‘students*/*teachers*/*exam agencies’, ‘medical research’ - ‘researchers’. The following of the review will show that existing publications on ChatGPT in healthcare can all find a proper categorization based on the proposed taxonomy.

**Figure 3:**
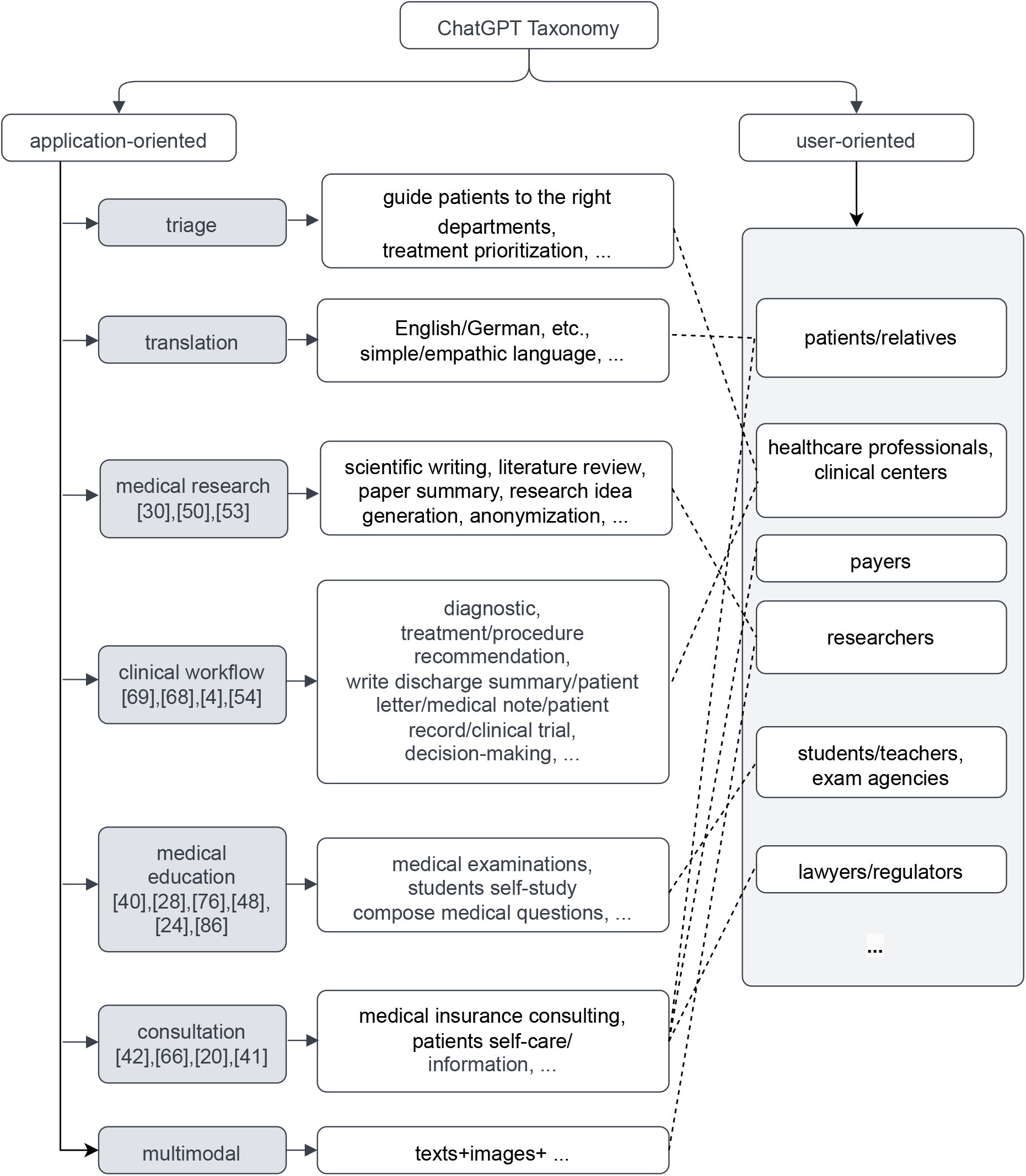
Application- and user-oriented Taxonomy used in the ChatGPT review. The references shown in the application boxes are the *Level 3* publications.

Besides the taxonomy, we further assign a *tag* (*Level 1* - *Level 3*) to the selected papers to indicate the depth and particularity of the papers on the ‘*ChatGPT in Healthcare*’ topic:

- *Level 1* : Generic comments about the potential applications of ChatGPT in healthcare or in a specific medical speciality and*/*or scenario;
- *Level 2* : Comments with one or more example use cases of ChatGPT in a specific medical speciality and*/*or scenario and moderate discussion about the correctness of ChatGPT’s answers;
- *Level 3* : Qualitative and quantitative evaluation of ChatGPT’s answers to a decent amount of speciality- and*/*or scenario-specific questions, with insightful discussion about the correctness and appropriateness of the ChatGPT’s answers.

Shortly prior to our review, a systematic review of ChatGPT in healthcare was published by Sallam, M. [91]. An inclusive taxonomy and a proper differentiation among the selected publications (*tag* : *Level 1, Level 2, Level 3*) is, however, lacking. We believe that the tag helps readers quickly filter and locate papers of interest. This review put more emphasis on *Level 3* papers, since they provide a clearer picture of the real capability of ChatGPT in different healthcare applications.

### 3.3 General Profile of *Level 1* and *Level 2* Papers

A list of *Level 1* and *Level 2* papers are summarized in Table 1. It is not unexpected that the majority of shortlisted papers fall into the *Level 1* and *Level 2* category. As seen from Table 1, most of *Level 1* and *Level 2* papers are short editorial comments or letters to the editor from multidisciplinary journals like *Nature* (https://www.nature.com/) and *Science* (https://www.science.org/), or speciality journals like nuclear medicine [3, 59], plastic surgery [79, 38], synthetic biology [106] and orthopaedic [81]. These publications usually deliver high-level comments about the potential impact and pitfalls of ChatGPT in healthcare [113], with a focus on medical publishing. Scientific journals are among the immediate stakeholders of the publishing industry on which ChatGPT will exert a significant impact. Thus, publishers introduce new regulations regarding the use of ChatGPT in scientific publications, in particular whether ChatGPT is eligible as an author and ChatGPT-generated texts are allowed. Answers from leading publishers like *Science* are in the negative [105, 16]. *Nature* also bans ChatGPT authorship but takes a slightly more tolerant stance regarding ChatGPT-generated content, subject to a clear statement of whether, how and to what extent ChatGPT contributed to the submitted manuscript [103, 27]. Main argument for the decision is that ChatGPT cannot properly source literature where its answers are derived from, causing unintentional plagiarism, nor can it take accountability as human authors do [105, 27]. The decision is echoed by the academic community [58, 97, 115, 66], agreeing that ChatGPT-generated content must be scrutinized by human experts before being used [58], as the generated content, such as references [105, 12, 40, 31] could be fabricated. Lee, J.Y. et al. [66] reiterated from a legal (e.g., copy-right law) perspective the inappropriateness of listing ChatGPT as an author, emphasizing that a non-human cannot take legal responsibilities and consequences. However, banning ChatGPT from scientific writing is not easily enforceable, since ChatGPT is trained to produce human-like texts that even scientists and specifically-trained AI detector sometimes fail to detect [29, 7]. In short, even though the prospect is promising [92, 25, 43, 102], new regulations and substantial improvements are needed before ChatGPT can be safely and widely used for scientific writing, publishing, or medical research in general [105]. The *scenario* column in Table 1 corresponds to the taxonomy categorization. If the article concerns healthcare or a medical speciality in general, it is categorized as ‘miscellaneous’. The *category* column indicates the type of the publications.

### 3.4 Reviews of *Level 3* Papers

*Level 3* papers feature extensive experiments conducted to assess the suitability of ChatGPT for a medical speciality or clinical scenario. For open-ended (OE) questions, human experts are usually involved to assess the appropriateness of the answers. To quantify the subjective assessments, a scoring criteria and scheme (e.g., 5-point, 6-point or 10-point Likert scale) is usually required. For multiple choice questions, it is desirable to not only quantify the accuracies but to evaluate whether the ‘justification’ given by ChatGPT and the choice are in congruence. When it comes to comparisons (with humans or other language models), statistical analysis is usually performed. As shown in Table 2, many of *Level 3* papers are still pre-prints (under review) at the time of writing this review. Most of current ChatGPT evaluations are on ‘medical education’ (medical exams in particular), which requires no ethical approval to conduct. Representative works include [36, 61], where the authors test ChatGPT in the US Medical Licensing Examination (USMLE). Even though the evaluations were carried out independently ([36] and [61] were published almost at the same time), similar results were reported, i.e., ChatGPT achieved only moderate passing performance. [36] further showed that ChatGPT outperformed two other language models, InstructGPT and GPT-3, in the exam. In both studies, ChatGPT was asked to give not only the answers but also the justifications, which were taken into consideration during evaluation (by physicians). [36] further found that ChatGPT performed better on fact-check questions than on complex ‘knowhow’ type questions. It is worthy of noting that the exam contains questions from different medical specialities. However, Mbakwe, A.B. et al. [74] raised concerns that ChatGPT, a language model, passing the exam indicates the flawness of the exam system ^1^. Besides USMLE, ChatGPT was also tested on the Chinese National Medical Licensing Examination [112] and the AHA BLS */* CLS Exams 2016 [32], on both of which ChatGPT failed to achieve passing scores.

ChatGPT achieved similar performance to students examinees on a Doctor of Veterinary Medicine (DVM) exam containing 288 parasitology exam questions. One major limitation of using ChatGPT in medical exams is that, current release of ChatGPT can only process text inputs, whereas some questions are diagram-*/*figure-based^2^. Such questions are either excluded or translated into text descriptions.

Besides the standard medical exams, ChatGPT achieved promising results on cancer-related questions [47, 53]. In [53], ChatGPT’s answers to common cancer myths and misconceptions were evaluated by expert reviewers and compared with the standard answers from the National Cancer Institute (NCI). Results showed that ChatGPT is able to achieve very high accuracies, showing that current ChatGPT is already a reliable source of cancer-related information for cancer patients [47]. Furthermore, [83] tested ChatGPT with 100 questions related to retina disease. The answers were evaluated based on a 5-point Likert scale by domain experts. It is found that ChatGPT answers with high accuracy on general questions, while the answers are less satisfactory, sometimes harmful, when it comes to treatment*/*prescription recommendations. On 85 multiple-choice questions concerning genetics*/*genomics, ChatGPT achieved similar performance to human respondents [28]. Interestingly, based on the test results, [28] also reached the conclusion that ChatGPT fares better on ‘memorization (fact-lookup)’ type questions than on those requiring critical thinking, similar to [83]. The performance of ChatGPT on these *question-answering* scenarios^3^ shows its potential for medical consultation and education.

A few studies evaluate the use of ChatGPT in medical research, particularly in scientific writing [67] and generating research questions [63] and systematic review topics [38]. In [67], the authors use ChatGPT to generate full abstracts, providing only the title and result sections of the abstracts from 50 real scientific publications. Even though previous studies [29] have shown that scientists can not tell apart abstracts generated by ChatGPT from those written by humans, [67] found that the two groups can simply be differentiated based on Grammarly scores. Discriminative features of ChatGPT-generated texts include mixed use of English dialects and language perfectness e.g., very few typos, more unique words, proper prepositions usage and no misuse of conjunction and comma. These characteristics can be captured by Grammarly scores. The finding indicates that Grammarly could potentially be adopted by scientific journals to enforce the ‘no-AI-generated-texts’ policy. In [63], the authors use ChatGPT to identify research questions in gastroenterology. The answers generated by ChatGPT proves to be highly relevant but lack depth and novelty. In [38], ChatGPT is used to generate systematic review topics in plastic surgery. Similar to [63], ChatGPT-generated research topics are generally not novel. The *version* column in Table 2 shows the version of ChatGPT used for evaluation. [63] found that newer versions of ChatGPT tend to have better performance on the same questions. In contrast to using ChatGPT directly for writing, which is expressly banned by many scientific journals, exploring new research ideas*/*topics with the assistance of ChatGPT faces less ethical issues. However, [63, 38] demonstrated that the current version of ChatGPT is not sufficiently qualified for such tasks. Humans still play dominating roles in ingenious and innovative research.

[87, 86, 4, 68] evaluate the application of ChatGPT in clinical workflow. In [87], ChatGPT is used to decide the appropriate imaging procedure (e.g., Mammography, MRI, US, etc.) for breast cancer screening and breast pain, given a description of the patients’ conditions. ChatGPT’s responses were evaluated against the corresponding American College of Radiology (ACR) appropriateness criteria. Results showed that ChatGPT achieved moderate overall results, and its performance is noticeably better for breast cancer screening than breast pain. The finding is in accordance with previous discussions that ChatGPT is already highly accurate on cancer-related information [47, 53]. The authors concluded that, even though ChatGPT showed impressive performance on the task, specialized AI tools are desired to support the clinical decision-making process more reliably. In a follow-up study [86], the authors tested ChatGPT with 36 clinical vignettes from the Merck Sharpe & Dohme (MSD), covering the entire clinical workflow (differential diagnosis, final diagnosis and subsequent clinical management of the patients). Overall, ChatGPT obtained a 71.8% accuracy in the test, and its performance on differential diagnosis is significantly lower than on final diagnosis. ChatGPT achieved the highest accuracy on a cancer vignette. The patients and their conditions in these vignettes are only hypothetical, which removes the ethical barrier to conduct the evaluation. In [4], ChatGPT is used to write patient clinic letters in 38 hypothetical clinical scenarios (e.g., basal cell carcinoma, malignant melanoma, etc.), where ChatGPT communicates the diagnosis results and treatment advice to the patients in a friendly and easily-understandable manner. The letters are evaluated from the perspective of factual correctness and humanness by clinicians, and ChatGPT achieved high scores on both criteria. In [68], ChatGPT is supplied with seven types of clinical decision support (CDS) alerts (e.g., pediatrics bronchiolitis, immunization, postoperative anesthesia nausea and vomiting, etc.) and asked to give suggestions. However, ChatGPT’s answers, even though highly relevant to the alerts, were not adequately acceptable by the standard of CDS experts.

## 4 Results

The following presents the answers to the four research questions (RQ1-RQ4) based on the discussion in Section 3.

### 4.1 Medical Applications of ChatGPT

According to Table 1, Table 2 and the taxonomy (Figure 3), it is straightforward to see that ChatGPT is mostly evaluated in medical education, consultation and research, as well as in various scenarios in the clinical workflow, such as diagnosis, decision-making and clinical documentation (patient letter, medical note, discharge summary, etc.). However, it is important to note these ‘applications’ are carried out in a ‘laboratory environment’, by providing ChatGPT question samples from standard medical exams (question banks), CSD alerts from Epic EHR or clinical vignettes from Merck Sharpe & Dohme (MSD), through its QA interface. None of the reviewed publications have reported an actual deployment of ChatGPT in clinical settings. Furthermore, due to the current strict policies on AI-generated content imposed by publishers, the unsolved ethical issues as well as its incapability in generating novel research topics, using ChatGPT for medical research remains experimental as well. For medical consultation, the fact that ChatGPT is already capable of providing highly accurate cancerrelated information can not be generalized to all medical specialities, since reliable sources of cancer information, such as the National Cancer Institute (NCI), are publicly accessible and could have already been part of ChatGPT’s training set. Its qualification as a medical consultant remains to be further evaluated.

### 4.2 Strengths and Limitations of ChatGPT in Healthcare

#### Strengths

The QA design of ChatGPT’s interface makes it easy to be integrated into existing clinical workflow, providing feedback in real-time. ChatGPT can not only give answers to specific questions but provide ‘justifications’ to its answers. Sometimes, ChatGPT’s ‘justifications’ and answers to open-ended question contain novel insights and perspectives, which might inspire novel research ideas. ChatGPT also shows superior performance in healthcare compared to other general large language models, such as InstructGPT, GPT-3.5.

#### Limitations

The current release of ChatGPT can only take input and give feedback in texts, so that ChatGPT cannot handle questions requiring the interpretation of images. ChatGPT is incapable of ‘reasoning’ like an expert system, and the ‘justifications’ provided by ChatGPT is merely a result of predicting the next words according to probability. It is possible that ChatGPT makes a correct choice, but gives completely nonsensical explanations. Accuracy of ChatGPT’s answers depends largely on the quality of its training data, and the information ChatGPT is trained on decides how ChatGPT would respond to a question. However, ChatGPT itself cannot distinguish between real and fake information fed into it, so that its answers could be highly misleading, biased and dangerous when it comes to healthcare. For example, one of the most concerning issues of current release of ChatGPT, as confirmed by the reviewed publications, is that it can ‘fabricate’ information and convey it in a persuasive tone. Therefore, its answers should always be fact-checked by human experts before adoption. Furthermore, ChatGPT’s answers, even if can be highly relevant, stay most of the time superficial and lack depth and novelty. Most importantly, ChatGPT is not fine-tuned for healthcare by design, and should not be used as such without specialization. Last but not least, the use of ChatGPT is not without barriers. Reformulating the prompt to the same question might change ChatGPT’s answer as well. Proper formulation of prompts is another factor to obtaining desirable answers from ChatGPT. Last but not least, ChatGPT is a proprietary product, and therefore feeding sensitive patient information into its interface in order to obtain a feedback might violate privacy regulations.

### 4.3 Research Gaps and Future Works

Prior to the deployment of any product in clinical settings, extensive evaluations of the product in a laboratory environment are required to identify the limitations and improve the product iteratively. Since ChatGPT was released no more than half a year ago, it has only been tested in a limited number of scenarios (Table 2). ChatGPT clearly is still at an experimental stage, and clinical deployment faces substantial unsolved technical and regulatory challenges. The *Level 3* publications provide a sound paradigm on how ChatGPT should continued to be evaluated in different specialities, for future works to follow. However, before further pursuing the direction, researchers should be aware that, even though these evaluations provide, at best, a general picture of ChatGPT’s capability in a medical speciality, little contribution to the improvement of the underlying language model is made. The limitations identified through these evaluations have also long been known in NLP research and are not specific to ChatGPT. Most importantly, whether or not ChatGPT has achieved good performance in an application scenario, it is unlikely that the ChatGPT with general knowledge will be clinically deployed in the future. Specialized AI models in healthcare, which the NLP community has long been working on, are more promising for practical and reliable clinical applications, compared to ChatGPT.

### 4.4 Categorization of Publications based on a Taxonomy

Finally, we have shown in our review that existing publications on ChatGPT in healthcare can be compactly grouped according to applications and target user groups. Thus, we come up with a application- and user-oriented taxonomy to categorize the selected publications, as discussed in Section 3.

## 5 Discussion and Conclusion

In this systematic review, we review published works (from Nov. 2022 to Mar. 2023) that used ChatGPT within the healthcare sector. In doing so, we extract publications from *PubMed* using the keyword ‘ChatGPT’ and propose a two-sided taxonomy (application-oriented and user-oriented) to categorize these publications, which we see as a building block for new publications on ChatGPT in healthcare. Even though the current taxonomy is already quite inclusive, it can be easily extended to emerging new applications or user groups. This first taxonomy is not limited to ChatGPT, rather it can also be applied to other (existing or upcoming) NLP models, like Bard from Google. On the one hand, the taxonomy helps interested readers to identify relevant works. On the other hand, it also helps identify areas where ChatGPT has not yet been applied to. An automatic processing of multimodal input, like text and images, is an exciting development for future healthcare. In example, Contrastive Language-Image Pre-Training (CLIP) [85], a neural network trained on large-scale *image-text* pairs, possesses both vision and language capabilities, and is therefore a promising research direction towards AI-assisted multimodal healthcare. In general, a physician takes also several sources of information into account when making diagnosis and treatment decisions, such as the written reports and image acquisitions from a patient. ChatGPT-4, a enhanced version of ChatGPT released recently, is able to analyse and summarize images and texts, as seen from a live demo given by its developers.

The barrier-free user interface, the ability to produce human-like texts and the breadth of its knowledge on a variety of topics are the key reasons why ChatGPT has amassed a phenomenally large user base shortly after its release. Besides the architectural design of the LLM, the immeasurable human efforts invested in training the LLM through reinforcement learning contribute greatly to its impressive performance in human-like conversations. Even though ChatGPT technically represents the productization of a NLP model by OpenAI, rather than a fundamental technological advance or breakthrough, it is undeniable that ChatGPT is a living embodiment of state-of-the-art NLP techniques. The efforts devoted to making the product a reality still greatly push forward the field as a whole. Speaking from the perspective of a tech product, existing publications on ChatGPT’s healthcare applications boil down to ‘reviews and testing of a new NLP product in healthcare’. However, the product is not intended for medical applications by design, and it is therefore not unexpected that most ‘test reports’ evaluated ChatGPT as ‘unqualified’ or ‘of merely passing grade’ for healthcare. However, the reported limitations (see Section 4) of ChatGPT are not specific to the product, but are applicable to language models in general, as discussed in Section 2. These limitations can mostly be addressed by improving the underlying language model through NLP innovations. Nevertheless, the fact that ChatGPT is monetized^4^ and therefore not (fully) open-sourced makes it difficult for the community to pinpoint the issues and come up with specific solutions for future improvement. In particular, the sources of datasets used for training the language model, which determine the type of questions and topics of the conversations ChatGPT can handle, remain unclear. As suggested by van Dis et al. [27], the community should invest in truly open LLMs that perform on par with proprietary NLP products like ChatGPT, in order to fully address these limitations. Currently, for healthcare applications, specialized AI models trained on biomedical datasets, such as BioGPT [70], are always more desirable than ChatGPT.

As discussed in this review (Section 3), these evaluation studies on ChatGPT’s performance in healthcare provide a general picture of the capability of the current release of ChatGPT. By and large, the training set and the underlying language model decide the quality (accuracy, unbiasedness, humanness, etc.) of the responses of an AI chat bot to certain questions. Therefore, this review concludes that healthcare researchers in particular should retract from the AI hype generated by the product and focus their attention on NLP research in general and developing*/*evaluating specialized language models for healthcare applications.

## Data Availability

All data produced in the present work are contained in the manuscript.

## Acknowledgments

This work was supported by the REACT-EU project KITE (Plattform für KITranslation Essen, EFRE-0801977, https://kite.ikim.nrw/) and the Cancer Research Center Cologne Essen (CCCE).

## Footnotes

1 ChatGPT does not fulfill the ‘USMLE Mission Statement’, but still passes the exam.

2 ChatGPT developers revealed that future versions of ChatGPT will have vision capabilities, and can comprehend images.

3 Exams are essentially also *question-answering*.

4 OpenAI has already introduced a subscription plan for ChatGPT (Plus).

